# Mathematical assessment of the role of intervention programs for malaria control

**DOI:** 10.1101/2023.12.18.23300185

**Authors:** Maame Akua Korsah, Stuart T. Johnston, Kathryn E. Tiedje, Karen P. Day, Jennifer A. Flegg, Camelia R. Walker

## Abstract

Malaria remains a global health problem despite the many attempts to control and eradicate it. There is an urgent need to understand the current transmission dynamics of malaria and to determine the interventions necessary to control malaria. In this paper, we seek to develop a fit-for-purpose mathematical model to assess the interventions needed to control malaria in an endemic setting. To achieve this, we formulate a malaria transmission model to analyse the spread of malaria in the presence of interventions. A sensitivity analysis of the model is performed to determine the relative impact of the model parameters on disease transmission. We explore how existing variations in the recruitment and management of intervention strategies affect malaria transmission. Results obtained from the study imply that the discontinuation of existing interventions has a significant effect on malaria prevalence. Thus, the maintenance of interventions is imperative for malaria elimination and eradication. In a scenario study aimed at assessing the impact of long-lasting insecticidal nets (LLINs), indoor residual spraying (IRS), and localized individual measures, our findings indicate that increased LLINs utilization and extended IRS coverage (with longer-lasting insecticides) cause a more pronounced reduction in symptomatic malaria prevalence compared to a reduced LLINs utilization and shorter IRS coverage. Additionally, our study demonstrates the impact of localized preventive measures in mitigating the spread of malaria when compared to the absence of interventions.

## 1 Introduction

Malaria remains a global health concern that threatens the lives of many children and adults each year. It has proven to be a persistent problem due to the highly adaptive nature of the *Plasmodium* spp. parasites and the female *Anopheles* mosquito vector [1, 2]. Over the past two decades, substantial headway has been made in reducing the global burden of malaria [3, 4]. These reductions are the result of political commitment, increased funding, and the wide-scale deployment of effective malaria control interventions targeting both the human host and the mosquito vector. However, in recent years progress has stalled and has even reversed in regions with moderate to high transmission. This rebound is particularly concerning for the most prevalent malaria parasite, *Plasmodium falciparum*, which is responsible for the majority of malaria-related deaths globally [5, 6]. To make matters worse, the COVID-19 pandemic demonstrated how even short-term disruptions in routine malaria interventions can impede progress in achieving elimination in malaria-endemic countries [7, 8].

Current malaria control strategies (or measures) include interventions that target the vector population and antimalarial therapeutic measures that target the human host population. Vector control strategies such as insecticide treated nets (ITNs)/long-lasting insecticidal nets (LLINs) and indoor residual spraying (IRS) with insecticides are key elements of current malaria control programs due to their effectiveness at interrupting transmission by reducing the population size of the mosquito vector [9]. Despite their historical success, these measures have drawbacks such as limited coverage (i.e., only target indoor transmission), being costly to implement and maintain, and potentially leading to the development of insecticide resistance in mosquitoes [10, 11, 12]. Therapeutic strategies, including the treatment of symptomatic infections with artemisinin-based combination therapy (ACTs) and targeted chemotherapy programs (e.g., intermittent preventive treatment (IPT) and seasonal malaria chemoprevention (SMC)) seek to reduce the number of infected human hosts, thereby reducing malaria morbidity and mortality, as well as onward transmission [9]. Unfortunately, these therapeutic measures have drawbacks like drug resistance and side effects, limiting long-term reliability [13, 14]. Given the limited budget available for malaria control, it becomes essential to optimize the allocation of resources and select interventions that provide the most significant impact. Thus, if malaria is to be eliminated in an endemic area, there is a need to adopt several strategic interventions simultaneously to avert both outdoor and indoor malaria transmission, and to curtail transmissions from the infectious human reservoir [12]. Mathematical modeling can help determine effective malaria interventions by providing valuable insights into complex disease dynamics and guiding decision-makers in the selection and implementation of the most cost-effective intervention strategies.

Mathematical modelling is effective in helping to tackle many epidemiological problems such as identifying disease determinants and controlling disease spread [15, 16]. Additionally, mathematical modelling has proven useful in the evaluation of malaria control programs and in assessing the transmission dynamics of infectious diseases amidst interventions [17, 18, 19]. In the study of malaria transmission, the *S*_*H*_ *E*_*H*_ *I*_*H*_ *R*_*H*_ − *S*_*M*_ *E*_*M*_ *I*_*M*_ model has been used widely as a simple yet practical approach to understanding the transmission patterns of malaria (and other vector-host infections), adding significantly to our knowledge of malaria [20, 21, 22, 23, 24, 25, 26]. For instance, Chitnis et al. and Osman et al. employed the *S*_*H*_ *E*_*H*_ *I*_*H*_ *R*_*H*_ − *S*_*M*_ *E*_*M*_ *I*_*M*_ model to examine the transmission dynamics of malaria in a human population [22, 25, 27]. They found that the rate of infection parameters in both humans and mosquitoes are the most influential parameters on the basic reproduction number, ℛ_0_ [25, 27]. Following the results obtained, the authors recommended reducing malaria prevalence with antimalarial treatment and reducing contact rates with IRS and ITNs/LLINs. Osman et al. also emphasised the importance of future research focusing on assessing the impact of interventions and conducting disease control analysis with the *S*_*H*_ *E*_*H*_ *I*_*H*_ *R*_*H*_ − *S*_*M*_ *E*_*M*_ *I*_*M*_ model and to date, a notable research gap remains in this space [25].

In this paper, we employ an extension of the *S*_*H*_ *E*_*H*_ *I*_*H*_ *R*_*H*_ − *S*_*M*_ *E*_*M*_ *I*_*M*_ model to consider the impact of interventions targeting the vector, such as IRS and ITNs/LLINs on the spread of malaria, specifically focusing on the transmission of *P. falciparum*, while factoring into the model the transmission characteristics of partial immune individuals. These are individuals who have acquired some level of protection after repeated exposure to malaria parasites but have not developed full immunity that would completely prevent infection. We assume that when these partially immune individuals become infected, they remain asymptomatic but can still transmit the malaria parasite, thus contributing to the continued transmission of malaria. This paper is structured as follows; in Section 2, a deterministic transmission model with separate transmission routes for non-immune and partially immune individuals is constructed. We formulate the basic reproduction number of the model and conduct a sensitivity analysis on the model parameters in Sections 3.1 and 3.2. In Section 4, we assess the impact of intervention strategies or measures on malaria transmission. Finally, we provide recommendations for improving malaria control programs, based on our results and discuss the implications of our findings in Section 5.

## 2 Model Formulation

Building on the disease transmission models of Yang et al. and Osman et al., we develop a malaria transmission model that takes into account transmission from both partial and non-immune infectious humans [17, 25]. We extend the *S*_*H*_ *E*_*H*_ *I*_*H*_ *R*_*H*_ − *S*_*M*_ *E*_*M*_ *I*_*M*_ model by splitting the susceptible and exposed human classes into two sub-classes each as similarly done by ul Rehman et al. [28], to capture the transmission properties of both non-immune and partially immune individuals. The human population therefore has six compartments, see Figure 1;

**Figure 1.**
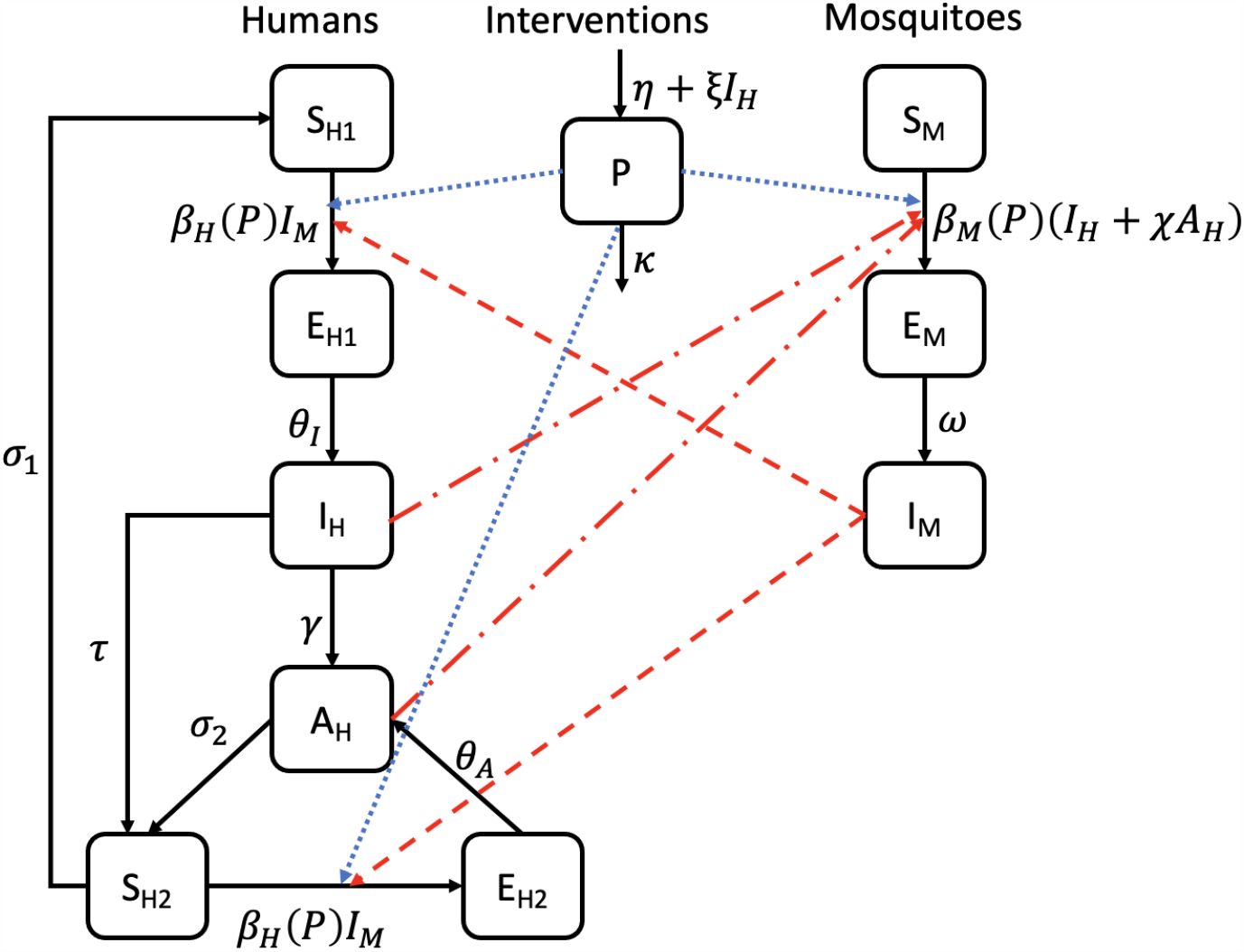
A compartmental diagram of the host-vector model. The red dash-dotted lines represent transmission from infectious humans to susceptible mosquitoes, the red dashed lines represent transmission from infectious mosquitoes to susceptible humans and the blue dotted lines symbolise the effect of intervention strategies on the transmission of malaria in both host and vector populations. Birth and death rates are not presented in this figure, though they are accounted for in the model. Refer to Table 1 for the definitions of the parameters of the model.

- *S*_*H*1_: non-immune, uninfected individuals susceptible to symptomatic infection,

**Table 1:** Definition of the model parameters.
- *S*_*H*2_: partially immune, uninfected individuals, susceptible to re-infection (asymptomatic),
- *E*_*H*1_: non-immune individuals in latent phase of symptomatic infection,
- *E*_*H*2_: partially immune individuals in latent phase of asymptomatic infection,
- *I*_*H*_ : symptomatic infectious individuals,
- *A*_*H*_ : asymptomatic infectious individuals.

| Parameter | Description | Baseline values | References |
| --- | --- | --- | --- |
| $\Lambda_H$ | influx rate of susceptible humans | 0.235 persons/day | [39] |
| $\Lambda_M$ | influx rate of susceptible mosquitoes | 26.7 mosquitoes/day | [39] |
| $\delta$ | mosquito biting rate | 1/ day | [39] |
| $\psi_H$ | probability of transmission from infectious mosquito to susceptible human during bite | 0.22 | [40, 39] |
| $\psi_M$ | probability of transmission from infectious human to susceptible mosquito during bite | 0.24 | [40, 39] |
| $\mu_H$ | natural death rate of humans | $4.5 \times 10^{-5}$ /day | [40, 39] |
| $\mu_M$ | natural death rate of mosquitoes | 0.0477 /day | [40, 39] |
| $\tau$ | treatment rate of symptomatic infectious humans | 0.08 /day | [39] |
| $\omega$ | latency rate in mosquitoes | 1/9 /day | [41, 42] |
| $\gamma$ | average untreated symptomatic infection duration rate in humans | 0.08 /day | [40, 39] |
| $\nu$ | disease induced mortality rate of humans | 0.08 /day | [40, 39] |
| $\chi$ | relative infectiousness of asymptomatic humans | 0.8 | - |
| $\eta$ | influx rate of intervention programs | 0.3/day | - |
| $\kappa$ | decay rate of intervention programs | 0.04/day | - |
| $\xi$ | growth rate of programs stimulated by symptomatic infections | 0.0025/day | - |
| $b$ | constant coefficient of $P$ in the human transmission rate function | 0.5 | - |
| $c$ | constant coefficient of $P$ in the vector transmission rate function | 0.4 | - |
| $\theta_I$ | latency rate in non-immune humans | 1/15 /day | [30] |
| $\theta_A$ | latency rate in partially immune humans | 1/20 /day | [39] |
| $\sigma_1$ | immunity waning rate of partially immune humans | 0.01 /day | [40, 39] |
| $\sigma_2$ | recovery rate from asymptomatic infectiousness | 0.01 /day | [40, 39] |

We divide the mosquito population into (*S*_*M*_) susceptible, (*E*_*M*_) exposed and (*I*_*M*_) infected mosquitoes.

We consider the impact of intervention strategies to provide valuable insights and evidence that can guide decision-making in reducing malaria transmission [17, 18]. We define intervention programs or strategies as measures that aim to lower the prevalence of malaria in an endemic region. To optimize the level of intervention programs needed for the elimination of malaria, we explore the effect of intervention programs (*P*) on the transmission of malaria. See Figure 1 for a schematic of the model structure. In the model formulation, we incorporate a constant influx of interventions with rate *η*. The influx of interventions is also influenced by the number of symptomatic infectious cases (*I*_*H*_) at a rate of *ξ* and the interventions decrease at rate *κ*. We assume that the availability and usage of intervention programs, *P*, affect disease trends by modulating the transmission rate.

The model in Figure 1, governed by the system of ODEs in Equation (1), can be used to assess the intervention programs necessary for the elimination of malaria in a geographic setting:

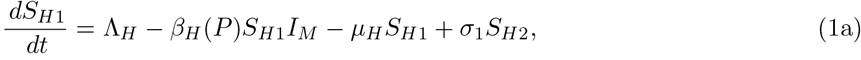

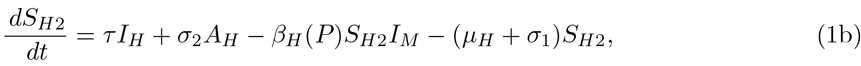

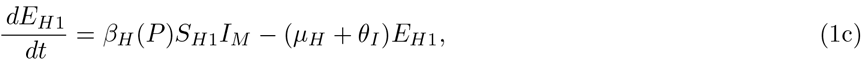

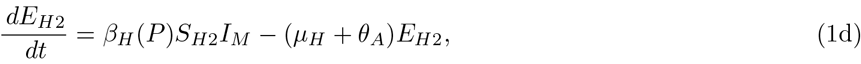

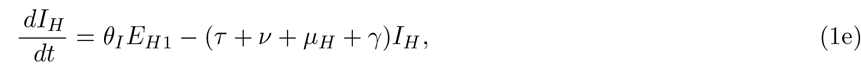

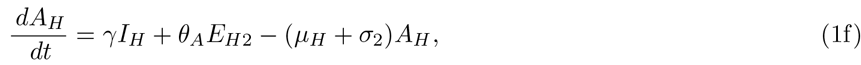

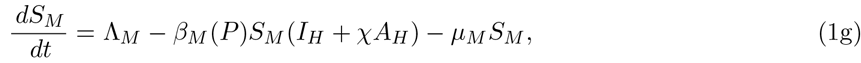

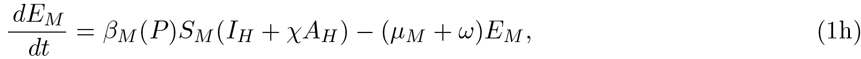

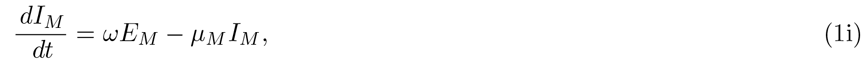

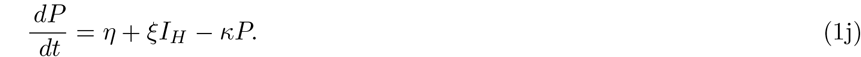

The model formulated in Equation (1) describes how susceptible individuals in *S*_*H*1_ have a population influx rate of Λ_*H*_ and are exposed to *P. falciparum* parasites by an infectious adult female mosquito during blood meals with a frequency-dependent transmission rate of *β*_*H*_ (*P*), moving individuals from *S*_*H*1_ into *E*_*H*1_. After a latent phase in *E*_*H*1_, individuals move into the symptomatic infectious class (*I*_*H*_) where they either self-recover at rate *γ* and move into the asymptomatic class (*A*_*H*_) or they are medically treated at rate *τ* with prescribed drugs such as ACTs [29, 30], and move into the *S*_*H*2_ compartment. The partially immune yet susceptible individuals in the *S*_*H*2_ compartment consist of treated individuals from the *I*_*H*_ class with noninfectious levels of the *Plasmodium* parasites due to treatments received, and asymptomatic persons with infection-induced immunity who transition into this class from the *A*_*H*_ class at rate *σ*_2_. Individuals here can either move back into *S*_*H*1_ by the loss of immunity at rate *σ*_1_ or move into *E*_*H*2_ by being re-exposed to *P. falciparum* parasites. From *E*_*H*2_ individuals become infectious with asymptomatic malaria (*A*_*H*_) at rate *θ*_*A*_. The model takes into account the human natural death rate in each class as *μ*_*H*_ as well as mortality due to clinical infection at rate *ν*.

In the mosquito population, susceptible mosquitoes are exposed to the malaria parasites at a frequency-dependent transmission rate of *β*_*M*_ (*P*) via transmission from both symptomatic and asymptomatic infectious humans and exposed mosquitoes become infectious at rate *ω*. Transmissions from asymptomatic infectious humans are scaled by a factor, *χ*, such that *χ* ∈ [0, 1) since asymptomatic humans infect mosquitoes at a lower rate than symptomatic infectious humans [31, 32, 33].

The intervention class of the model, *P*, affects the transmission rate functions of the model. Thus, interventions considered here can capture the impact of vector control strategies (IRS and ITNs/LLINs), intermittent preventive treatment (IPTs) with antimalarials, individual measures (environmental preventive measures) such as clearing mosquito breeding sites and/or reducing exposure to mosquitoes (e.g., personal repellents, insect coils and room sprays), and other approaches that can interfere with malaria transmission [34, 35]. The intervention class modelled here does not consider other malaria therapeutic measures like ACTs as they affect other aspects of the model.

We further assume that:

(H1) All parameters are non-negative.

(H2) The frequency-dependent transmission rate functions are defined as decreasing functions of the intervention programs *P* ;

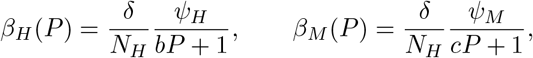

where *b* and *c* are positive-valued constants, *N*_*H*_ is the total human population, (*N*_*H*_ = *S*_*H*1_ + *S*_*H*2_ + *E*_*H*1_ + *E*_*H*2_ + *I*_*H*_ + *A*_*H*_), *δ* is the biting rate, and *ψ*_*H*_ and *ψ*_*M*_ are the infection success probabilities in humans and mosquitoes respectively.

(H3) The transmission rate functions are decreasing functions of *P* (i.e. 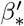(*P*) *<* 0) and *P* takes values in [0, *P*_*max*_] where

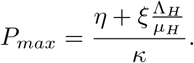

## 3 Model Analysis

### 3.1 Formulation of Basic Reproduction Number, *ℛ* _0_

To better understand the proposed framework, we evaluate the basic reproduction number which quantifies new cases generated near the disease-free equilibrium (DFE). The basic reproduction number is formulated using the next generation method established by Diekmann et al., and Van den Driessche and Watmough [36, 37, 38]. Refer to Appendix A for the detailed derivation of the basic reproduction number of the model.

The basic reproduction number of the model can be formulated as,

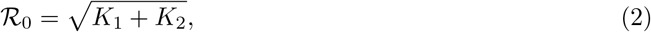

where

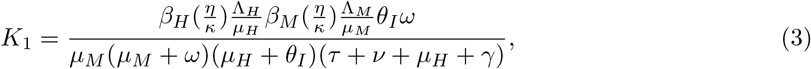

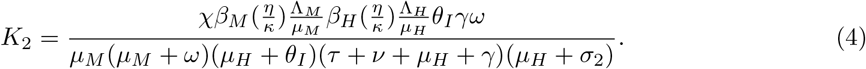

From Equation (2), we identify two transmission links;

- *K*_1_, which represents transmission from individuals in *I*_*H*_, and
- *K*_2_, which reflects transmission from self-recovered individuals in *A*_*H*_.

Transmission from infected mosquitoes is accounted for in each transmission pathway. In the absence of intervention programs (*P* = 0), the basic reproduction number becomes:

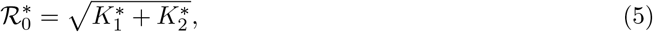

where

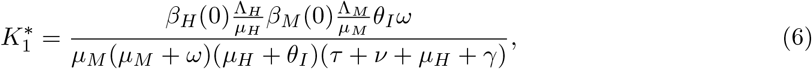

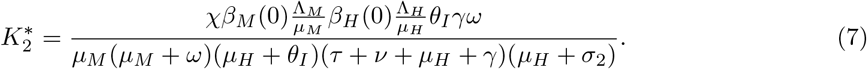

Thus,

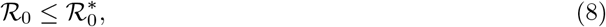

since the transmission rate functions are decreasing functions of *P*, reflecting the impact of intervention programs to reduce the spread of malaria.

We note that ℛ_0_ does not account for transmission from re-infections, which is due to the fact that there are no individuals with partial immunity at the DFE.

### 3.2 Sensitivity Analysis on ℛ_0_

We conduct a sensitivity analysis on ℛ_0_ to obtain qualitative information on how the model parameters affect ℛ_0_ by employing the normalized forward index, *ζ* of ℛ_0_ for a parameter *k*, [43] as

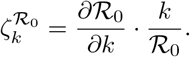

We compare the sensitivity index of the parameters on ℛ_0_ and 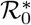 in Equations (2) and (5), and observe that the normalised forward index, *ζ* of both ℛ_0_ and 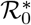 is the same for all parameters except for *b, c, η* and *κ*, which are parameter related to the interventions class *P* as 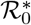 is formulated in the absence of intervention strategies. The results obtained are presented in Table 2.

**Table 2:** Sensitivity index of the model parameters on ℛ_0_ and 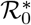.

| Parameters | Sensitivity index on $\mathcal{R}_0$ and $\mathcal{R}_0^*$ |
| --- | --- |
| $\delta$ | 1 |
| $\psi_H$ | 0.5 |
| $\psi_M$ | 0.5 |
| $\Lambda_M$ | 0.5 |
| $\omega$ | $\frac{\mu_M}{2(\mu_M + \omega)} > 0$ |
| $\chi$ | $\frac{\chi\gamma}{2(1 + \chi\gamma)} > 0$ |
| $\theta_I$ | $\frac{\mu_H}{2(\mu_H + \theta_I)} > 0$ |
| $\gamma$ | $\frac{\gamma[\chi(\tau + \nu + \mu_H) - 1]}{2(\tau + \nu + \mu_H + \gamma)(1 + \chi\gamma)} > 0$ |
| $\mu_M$ | $-\frac{3\mu_M + 2\omega}{2(\mu_M + \omega)} < 0$ |
| $\tau$ | $-\frac{\tau}{2(\tau + \nu + \mu_H + \gamma)} < 0$ |
| $\nu$ | $-\frac{\nu}{2(\tau + \nu + \mu_H + \gamma)} < 0$ |
| $\sigma_2$ | $-\frac{\sigma_2}{2(1 + \mu_H + \sigma_2)} < 0$ |
| Interventional Parameters | Sensitivity index on $\mathcal{R}_0$ |
| $\kappa$ | $\frac{\eta[2bc\eta + (b+c)\kappa]}{2(b\eta + \kappa)(c\eta + \kappa)} > 0$ |
| $b$ | $-\frac{b\eta}{2(b\eta + \kappa)} < 0$ |
| $c$ | $-\frac{c\eta}{2(c\eta + \kappa)} < 0$ |
| $\eta$ | $-\frac{\eta[2bc\eta + (b+c)\kappa]}{2(b\eta + \kappa)(c\eta + \kappa)} < 0$ |

Based on Table 2, we deduce that parameters such as the mosquito biting rate (*δ*), infection success probabilities (*ψ*_*H*_, *ψ*_*M*_) and decay rate of intervention programs (*κ*), which have positive indices contribute to the initial spread of malaria (in that as the parameter increases, ℛ_0_ increases). In contrast the treatment rate of infected persons (*τ*), interventions recruitment/funding rate (*η*), and mosquito death rate (*μ*_*M*_) parameters with negative indices reduce ℛ_0_. Some model parameters are not included in Table 2, such as *σ*_1_, *θ*_*A*_, and *ξ*, since they do not exert a direct influence on ℛ_0_, see Equation (2). Note that we exclude the human birth and death rates from the ℛ_0_ sensitivity analysis, as these factors are not directly adjustable in the context of malaria control strategies. The focus is on parameters that are amenable to intervention, which is more pertinent for policy considerations.

## 4 Numerical Results

### 4.1 Local Sensitivity Analysis on ℛ_0_

We substantiate the parameter sensitivity results in Table 2 by plotting ℛ_0_ and 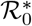 as a function of individual parameters in Figure 2. As expected, parameters with positive indices exhibit a positive impact on both ℛ_0_ and 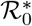, while conversely, parameters with negative indices have a negative effect. The difference in impact between ℛ_0_ and 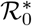 is clear from Figure 2, and indicates that the absence of intervention programs leads to an increase in the basic reproduction number, which supports the findings from Equation (8).

**Figure 2.**
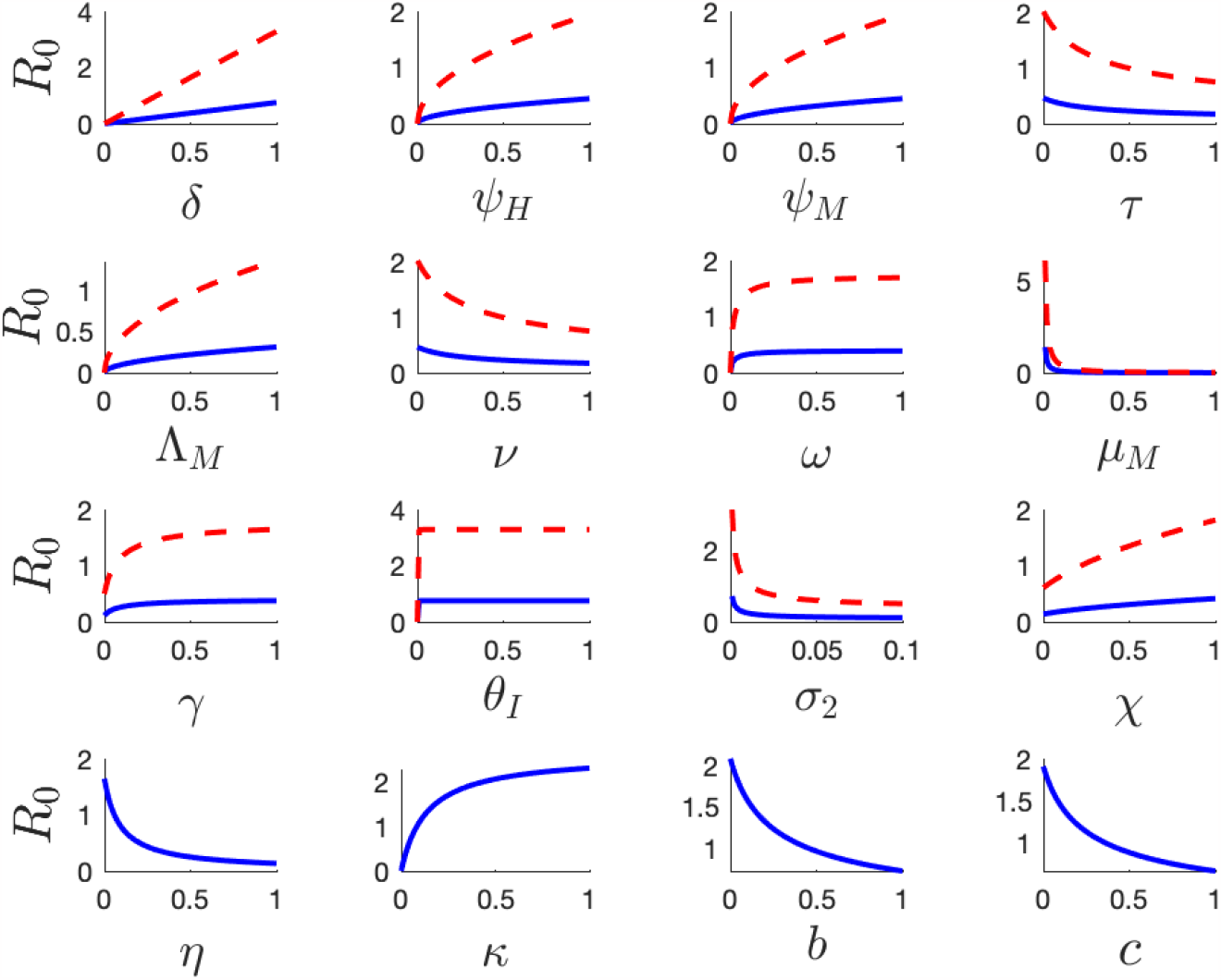
Plots from a local sensitivity (see Equations (2) and (5)) analysis conducted on the model. In these plots, the blue solid lines illustrate the variations in ℛ_0_, as individual parameters change, whereas the red dashed curves illustrate the variations in ^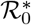^ with respect to the specific parameter being analyzed, with all other parameters maintained at their baseline values as detailed in Table 1.

### 4.2 Impact of Variation in Interventions on Malaria

In Figure 3, we present the outcomes of a sensitivity analysis to investigate the combined effect of changes in *η* and *κ* on ℛ_0_. We observe that the intervention decay rate, *κ*, has an increasing effect on ℛ_0_ whereas the recruitment rate of interventions, *η*, has an inhibitory effect on ℛ_0_ which is consistent with ℛ_0_ increasing as interventions (*P*) increases.

**Figure 3.**
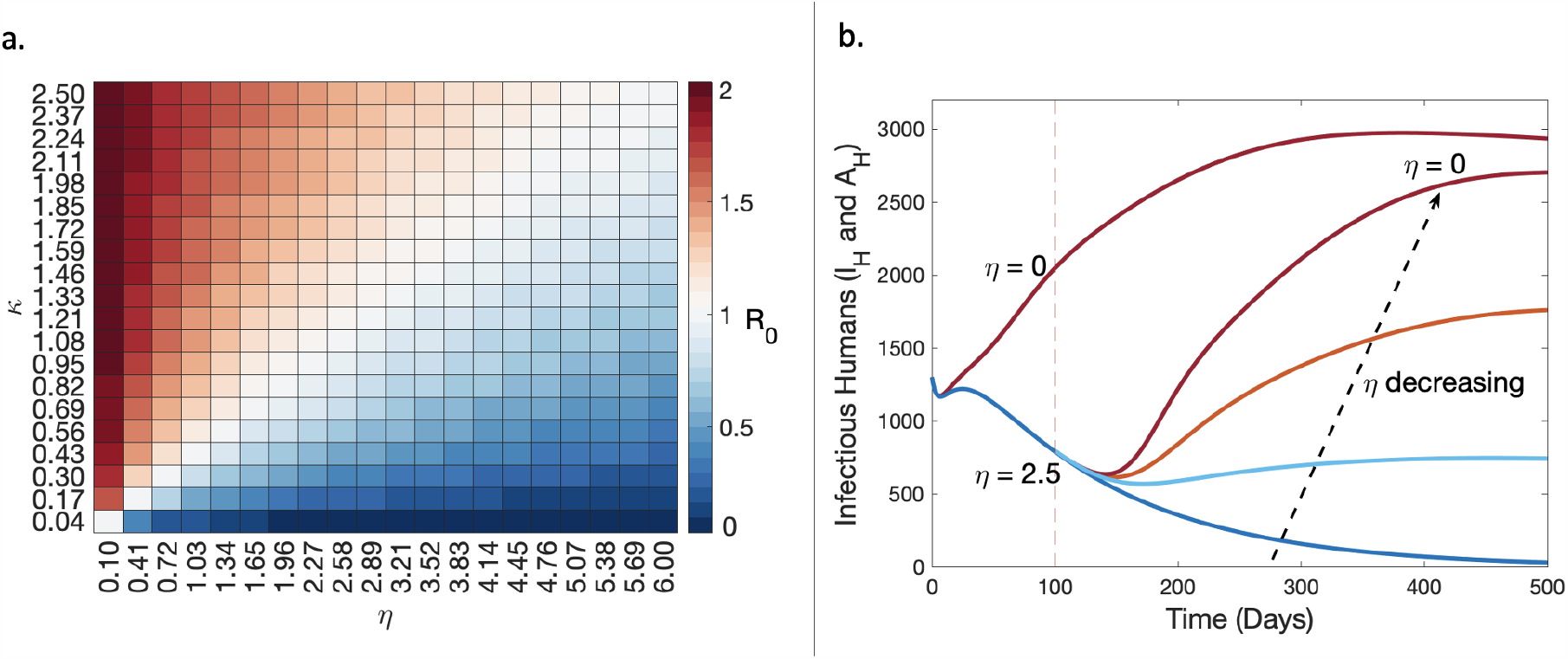
**a**. A sensitivity analysis of ℛ_0_ on the intervention parameters *κ* and *η*. The colour bar represents ℛ_0_ values ranging from 0 (blue) indicating a negative growth of the disease to 2 (red) indicating a positive initial disease growth. The subplot **a**. is derived from the solution to Equation (2). **b**. Simulation results comparing the dynamics of infectious human cases from a steady declining situation (dark blue line), as the recruitment rate of intervention programs is decreased at 100 days compared to having *η* = 0 from the start. Four decreasing scenarios of *η* are considered at 100 days from *η* = 2.5 (dark blue line) to *η* = 0 (dark red line) with *κ* = 0.17 in all scenarios. Results in subplot **b**. are obtained by solving Equation (1) and recording changes in Equations (1e) and (1f) as *η* varies.

From Figure 3a, we have identified the level of recruitment and decay of intervention measures necessary to prevent malaria outbreaks given the dynamics of transmission in an endemic setting (e.g. that push ℛ_0_ *<* 1). The white region of Figure 3a represents the level required to control initial malaria transmission, where ℛ_0_ drops to 1. The blue region represents the region of the parameter space where malaria is suppressed but achieves a more substantial reduction in ℛ_0_ than necessary to suppress outbreaks.

We also perform numerical simulations of the model to investigate how variations in intervention strategies resulting from decreasing intervention funds and the discontinuation of established strategies affects the number of infectious cases in the human population before and after disease elimination. Here we classify infectious cases, *I*, as the sum of symptomatic and asymptomatic infectious humans, that is, *I* = *I*_*H*_ + *A*_*H*_. We consider a situation where a diminishing trend in malaria cases results in reduced funds for antimalarial interventions [44, 45]. This behaviour of funding agencies is factored into the model by reducing the recruitment rate of interventions, *η*. It is important to clarify that the concept of funding in this context represents the influx into *P*, rather than a specific dollar value. Figure 3b presents the simulation results of the number of infectious humans under different varying scenarios of *η*. We observe that as funding for malaria elimination programs decreases, infectious cases increase. This result was generated under the assumption that when there is a 40% decrease in infectious cases, stakeholders will consider decreasing funding rates. The scenario where *η* = 0 after the initial disease decline, leads to a higher increase in infectious cases, closer to the “no control” situation with *η* = 0 from the start. In fact, this scenario with *η* = 0 after the initial decline will ultimately hit the same steady state as the “no control” situation. However, maintaining *η* = 2.5 sees malaria approach elimination. Thus we can infer from the results of Figure 3b that in order to sustain a diminishing trend in infectious malaria cases, it is likely necessary to either raise or maintain the funding rates for intervention recruitment strategies.

To explore the post-elimination prospects of malaria in endemic regions, we conducted numerical experiments using the baseline parameter values in Table 1, setting *δ* = 3, *ψ*_*H*_ = 0.5, and *ψ*_*M*_ = 0.5, until the time point where the number of infectious individuals, denoted as *I*, falls below a specified threshold *ϵ*. Using the same parameter values and compartmental dynamics observed at that particular point in time (i.e. we set *I*_*H*_ (0) = *I*_*H*_ (at elimination) + 1 and maintained the values of all other compartments), we then introduced a single symptomatic infectious human and varied the decay rate of the intervention strategies, *κ* (as per Figure 4a). In reality, malaria elimination is when *I* = 0, how-ever in practice we set malaria elimination status at a threshold of *ϵ* = 0.3 *>* 0 since we are employing a continuum model. By varying the value of *κ* in Figure 4a, we observe an increase in infectious cases as the decay rates of interventions increases. We thus infer that the discontinuation of intervention programs after elimination could lead to a reemergence of malaria. Setting *κ* = 0, however, results in an unperturbed malaria elimination state even with the introduction of an infectious case as strategies to control the spread of malaria are still available. Our results suggest that maintenance of established intervention strategies is, therefore, necessary to maintain elimination status.

**Figure 4.**
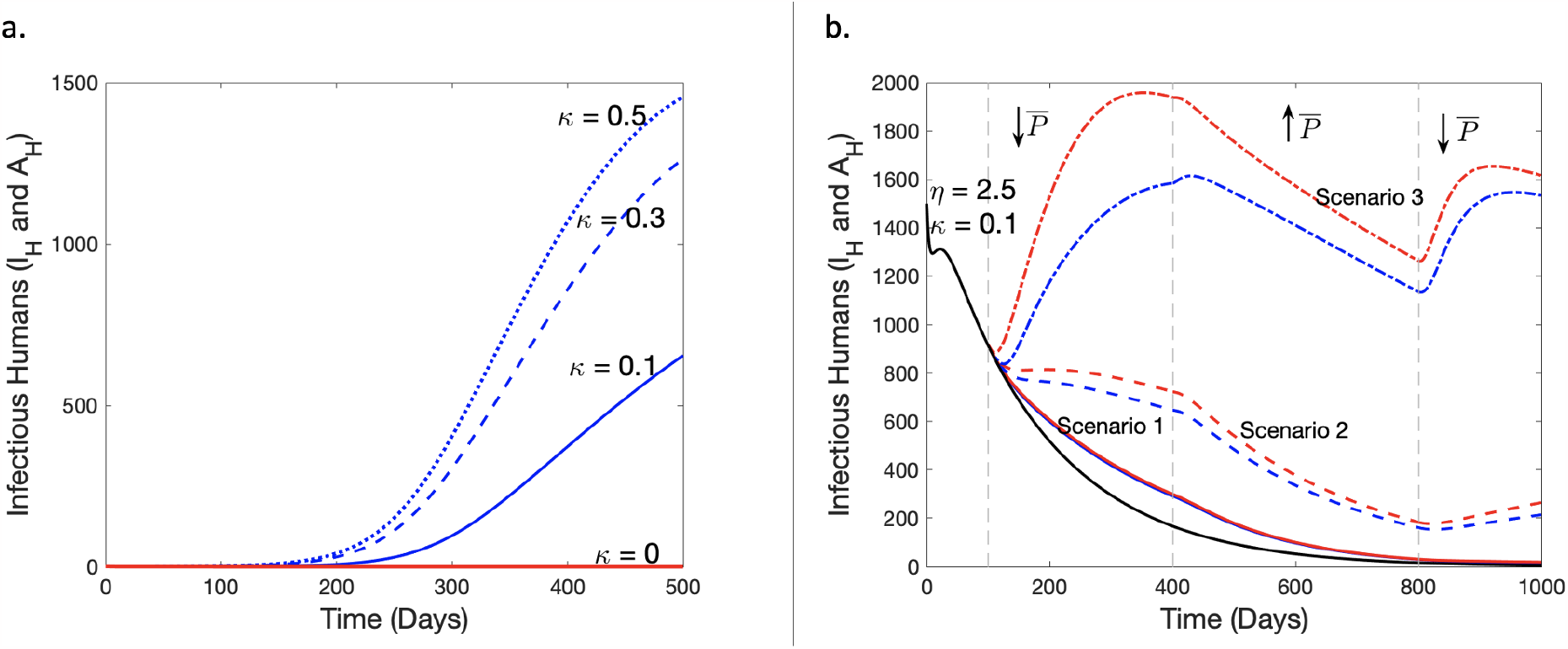
Simulation results exploring how variations in interventions affect infectious trends. **a**. After-elimination scenarios on how the management (decay) of intervention strategies can affect infectious human cases in the first 500 days post-elimination. After elimination, we set *η* = 0 and consider four scenarios of *κ*; *κ* = 0 (red solid line), *κ* = 0.1 (blue solid line), *κ* = 0.3 (blue dashed line) and *κ* = 0.5 (blue dotted line). **b**. The impact of time-varying intervention strategies on malaria-infectious human cases. The solid black line represents the ideal (baseline) scenario with a constant supply and decay rate of interventions, the blue line represents results from variations in the recruitment rate of intervention strategies, *η*, while the red lines represent simulation results from a corresponding change in the decay rate of interventions, *κ*. The changes are chosen such that 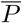, (i.e. *P* at the DFE) for each change in *η* and *κ* is equal. We consider three scenarios of unsteady intervention strategies from day 100 and explore further variations in the supply and decay rate of interventions from days 400 and 800. These days are marked by the vertical grey dashed lines. Scenario 1 is marked by the solid red and blue lines, Scenario 2 by the dashed lines, and Scenario 3 by the dash-dotted lines (specific details are discussed in the main text). Solution of both subplots **a**. and **b**. are obtained by solving the model system in Equation (1) and recording the specific changes in Equations (1e) and (1f).

In light of the current erratic trends in malaria cases globally, we investigate the dynamics of infectious cases resulting from a changing supply of funds for malaria intervention programs and the temperamental utilization of interventions by exploring variations in *η* (∆*η*) and *κ* (∆*κ*) such that 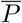, the *P* at the DFE, for each change in *η* and *κ* is equal (i.e. 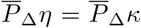, where 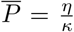). In each scenario, a percentage change in 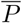 is achieved via a modification of the parameter *η* or *κ*. From Figure 4b, we observe three scenarios (specific details below) of trends in malaria cases resulting from an initial decrease in 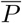 after the baseline scenario at day 100. This is followed by a substantial increase in 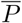 from day 400, aimed at rectifying possible increasing trends of infectious cases. Finally, 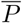 is decreased after day 800 by a percentage less than the initial decrease at day 100. This is done to incorporate positive but deficient human behaviour targeted towards malaria elimination into our investigation of the unsteady trends in malaria cases. We discuss the details of variations considered in all three scenarios below.

- Scenario 1 (solid lines in red and blue) – After 100 days of the baseline simulation, we decreased 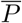 by 30% and observed similar results for the variations in both *κ* (red) and *η* (blue), that is a general reduction in malaria cases. After day 400, we increase 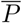 by 50% and observe human infectious cases fall closer to elimination levels. With the number of infectious cases nearing elimination, a subsequent 20% decrease in 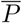 at day 800 results in further declines as the infectious cases present are not enough to cause a rise in cases.
- Scenario 2 (dashed lines in red and blue) – After day 100, we decrease 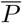 by 55% which results in a higher increase in cases for the corresponding change in *κ* (red) than in *η* (blue) by day 400. We then increase 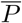 by 75% and observe a similar reduction trend in both *η* and *κ* from day 400 to 800. We finally reduce 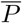 by 35% after day 800 to observe an increase in both at day 1000.
- Scenario 3 (dash-dotted lines in red and blue) – 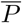 is cut by 80% after day 100 which leads to a substantial rise in malaria cases by day 400. We continue by increasing 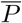 by 90% and observe a sharp decline in malaria cases for the change in *η* (blue) compared to *κ* (red) by day 800. We finally cut 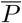 by 50% after day 800 and observe a climb in cases.

In all three scenarios presented in Figure 4b, we show that while the inconsistent supply of funds for intervention strategies (results for changing *η* in blue) has a notable impact on infectious cases, the unsteady maintenance of interventions (results for changing *κ* in red) has a more pronounced impact on infection trends.

### 4.3 IRS, LLINs and Individual Interventions Scenarios

We conduct a scenario study to investigate how IRS, LLINs, and individual (preventive) measures such as clearing mosquito breeding sites and reducing mosquito exposure [34, 35], affect the patterns of symptomatic malaria infections. To this effect, the model parameters, *η, κ, ξ, b* and *c*, which are associated with the intervention class were adjusted to reflect the effectiveness of the selected interventions. Drawing from existing literature, we incorporate into our scenario analysis the intervention’s implementation, durability and efficiency. Notably, LLINs exhibit an efficiency rate of approximately 77%, decreasing malaria prevalence by about 77%, and have a lifespan of three years [46, 47, 48]. Based on this information, we set *b* and *c* at 0.8 to mimic a similarly high efficacy of LLINs. Note that *b* and *c* are the constant coefficients of *P* in the human and vector transmission rate functions respectively, modelled to capture the effect of intervention strategies on the transmission rate functions. We assume a 0.5/day influx rate of LLINs with its usage growing at a rate of 0.08/day/symptomatic case as stimulated by the number of symptomatic cases (see LLINs column of Table 3). In the study, we also examine how the extent of LLINs usage (and the duration of IRS coverage) can influence symptomatic infections. Since the duration of LLINs exceeds the timeline of this study (a year), we model the *κ* values of LLINs to majorly capture the extent of LLINs usage. We assume a 90% usage to represent a high level of usage of bed nets and 40% percent as low usage. This percentage difference was factored into the choice of *κ* values since the ratio of 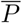 for high usage to 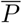 for low usage is 9:4 (refer to LLINs column of Table 3 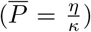). IRS, on the other hand, demonstrates high varying efficacy depending on coverage levels, and a duration ranging from 5 to 8 months contingent upon the specific chemical employed [49, 50, 51, 52, 53, 54]. In the IRS scenario, we assume a high coverage spraying is done at the start of the study that on average, decays in 5 months for the short duration case and 8 months for the long duration case. These durations were translated into the *κ* values found in the IRS column of Table 3. The parameters *b* and *c* we set at 0.85 to depict a high efficacy rate while *η* and *ξ* were set to 0 in line with the assumption that IRS intervention will not be administered again for the period of 1 year considered. Conversely, we assume relatively lower efficiency and durability for the localized individual interventions, see Individual Measures column of Table 3. However, we consider a higher usage of these individual measures when infectious cases are on the rise, acknowledging the adaptable nature of human behavior in response to changing disease dynamics [34]. Assuming a yearly recurrent implementation of these interventions, we run simulations for 365 days to assess and compare the effects of these three strategies against the baseline scenario, where no interventions are employed. In the baseline scenario, we assume that no intervention measures are active during the entire study period, and we set all related parameters to zero (see No Interventions column of Table 3). In Figure 5, we present a summary of the results of this study and provide the intervention parameter values utilised in the simulation study in Table 3.

**Table 3:** Intervention parameter values utilised for the simulation results in Figure 5.

| Parameters | No Interventions | Individual Measures | IRS |  | LLINs |  |
| --- | --- | --- | --- | --- | --- | --- |
|  |  |  | Long Duration | Short Duration | High usage | Low usage |
| $\eta$ | 0/day | 0.005/day | 0/day | 0/day | 0.5/day | 0.5/day |
| $\xi$ | 0/day | 0.001/day | 0/day | 0/day | 0.08/day | 0.08/day |
| $\kappa$ | 0/day | 0.15/day | 0.018/day | 0.03/day | 0.2/day | 0.45/day |
| $b$ & $c$ | 0 | 0.4 | 0.85 | 0.85 | 0.8 | 0.8 |

**Figure 5.**
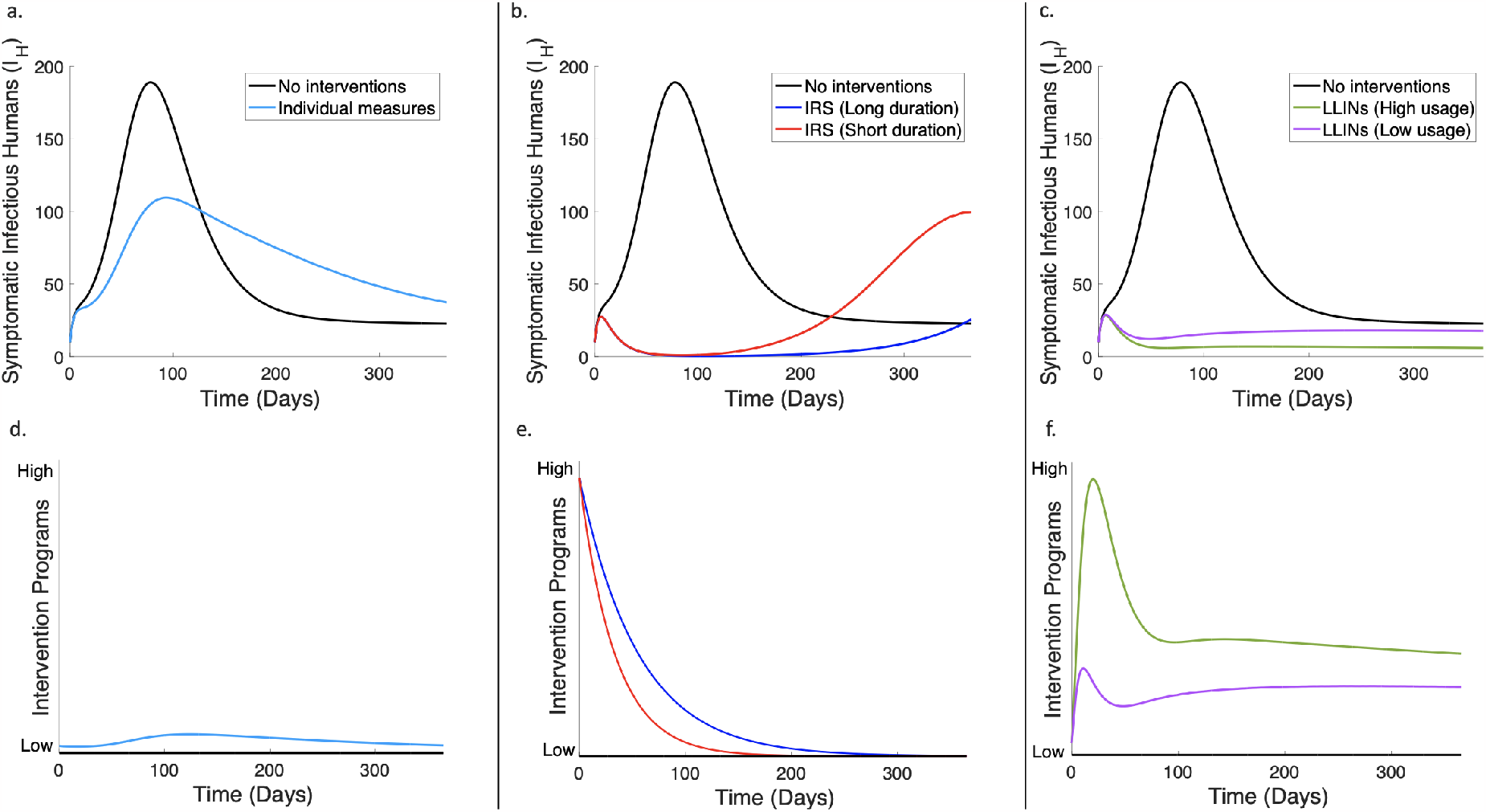
Simulation results capturing the impact of IRS, LLINs, and individual preventive measures on symptomatic infectious humans over a period of 365 days. **a**. Results comparing the impacts of individual measures on symptomatic malaria cases with the baseline when no interventions are implemented. **b**. Results comparing the impacts of IRS (long and short duration) with the baseline of no intervention measures on symptomatic infectious cases. **c**. Results comparing the impacts of LLINs (high and low usage) with no interventions on symptomatic infectious cases. **d.–f**. Graphical representation of the levels of interventions considered in the simulation studies in **a.–c**. respectively.

Figure 5 captures the impact of IRS, LLINs and individual measures on symptomatic infectious humans. In Figures 5a. and 5d. we observe that individual preventive measures that exhibit a relatively low impact amongst the three scenarios, can reduce symptomatic infections during the period of heightened disease activity. In an extended timeframe, beyond 365 days, we observe a declining trend in cases when using these individual measures, which falls below the baseline scenario. In Figures 5b. and 5e., our observations indicate that implementation of IRS leads to a significant reduction in malaria infection cases, within its designated effective duration. After IRS efficacy wanes there is an upsurge in the number of infectious cases. Specifically, within the 365 day period the 5-month IRS scenario has a peak prevalance lower than the peak of the baseline scenario, while in the 8-month IRS scenario has an even lower peak. In Figures 5c. and 5f., high usage of LLINs has greater impact on symptomatic infectious cases. Conversely, the scenario involving low LLINs usage initially demonstrates a decline in the first 50 days. However, as the number of cases reduces, the utilization of bed nets decreases further, leading to a resurgence in cases approaching the baseline scenario, as observed in Figures 5c. and 5f.

Taking into account all the interventions considered in the scenario study, our results suggest that the extensive utilization of LLINs and IRS within their designated effective durations has the potential to curb endemic malaria trends effectively. Nonetheless, when LLINs, IRS, and other highly effective interventions are unavailable, our modelling reveals that implementing individual preventive measures is better than adopting no interventions in the long run.

## 5 Discussion

In this paper, an extension of the *SEIR*− *SEI* host-vector model is employed to study the impact of malaria intervention programs. Our work is targeted towards understanding the current trends of malaria cases in response to malaria interventions as well as assessing the interventions necessary for malaria control and elimination. The extended model is analysed to formulate the basic reproduction number, consistent with previous studies [17, 20, 25, 55, 56, 57]. In our assessment of the basic reproduction number of the model, we identified two transmission pathways that can assist decision-making in the prevention of malaria outbreaks, which we termed as *K*_1_ and *K*_2_ (Equation (2)). *K*_1_ represents transmissions from individuals in the symptomatic infectious human class, *I*_*H*_, whereas *K*_2_ considers transmissions from self-recovered individuals in the asymptomatic infectious human class, *A*_*H*_. Thus *K*_1_ transmissions can be reduced by employing vector control strategies, intermittent preventive treatments of malaria in infants, pregnant people, and children, as well as the RTS,S/AS01 (RTS,S) vaccine recommended by the WHO for the prevention of *P. falciparum* malaria in children. *K*_2_ transmissions on the other hand, can be reduced with strategies like mass screening and treatment (MSAT), focal screening and treatment (FSAT), and mass drug administration (MDA) that typically focuses on asymptomatic infections as well as through educative campaigns promoting the clinical treatment of malaria cases with strategies like mass fever treatment (MFT) to reduce the number of symptomatic infections that go untreated [58, 59, 60, 9].

A sensitivity analysis conducted on the model demonstrates that several parameters such as the mosquito biting rate (*δ*), infection success rates (*ψ*_*H*_, *ψ*_*M*_) and decay rate of intervention programs (*κ*) have a positive influence on ℛ_0_ whereas parameters like the treatment rate of infected persons (*τ*), interventions recruitment rate (*η*), and mosquito death rate (*μ*_*M*_) have a negative impact on ℛ_0_. These results are in consensus with the findings of existing literature [25, 55, 61].

Our study on the impact of variations in preventive intervention strategies shows that reducing funds for malaria interventions in response to a decline in the number of malaria cases may result in the resurgence of malaria. These results reflect the current rising trends of malaria cases after the gradual decline in malaria cases from 2017-2019, as funding for malaria intervention programs was reduced in order to support the control of COVID-19 outbreaks [62, 63, 64]. Learning from the current malaria situation, our modelling results suggest that funding for intervention strategies should be maintained and increased when feasible but not decreased. We also discovered that the maintenance of intervention programs is necessary to maintain a malaria elimination status. Thus regions that have successfully eliminated malaria from their settings must endeavour to maintain malaria strategies longer term, such as detecting and treating new malaria infections among migrants, to prevent the re-emergence of malaria cases [65, 66]. Results derived from our intervention scenario study reveal that implementing IRS with an extended effective duration and promoting the extensive utilization of LLINs represent promising strategies for mitigating symptomatic malaria infections. These findings align with empirical results from previous literature [51, 67, 68, 69]. Additionally, the study illuminates the potential utility of individual preventive measures, albeit with a relatively low impact. These measures can be considered either in conjunction with other interventions or as a viable option when more potent interventions are not available.

Moving forward, an expansion of this work is recommended to calibrate the model to data from a malaria endemic setting while inferring parameters for the interventions used in the setting. Factors such as age structure, seasonality, and migration, which are major determinants of malaria trends in endemic regions, were not factored in the model and could be considered in future extensions. Additionally, the interventions class of the model did not explicitly consider the dynamics of strategies such as RTS,S vaccine roll-out, the use of larvicides, MSAT, and the development of better healthcare systems in endemic areas. While these limitations may affect the application of results presented in the study, the study provides an overview of the transmission characteristics of a typical endemic area and thus can be adapted to a specific setting by including additional characteristics of that setting into the model.

Several concerns remain unresolved in relation to the behavioural trends of the *Plasmodium* parasites such as their heterogeneity and drug resistance which hinder the elimination of malaria. However, the results of this study suggest that the continuous maintenance of established intervention strategies in endemic areas can provide progress towards malaria elimination. While variations in the implementation of interventions may occur due to economic constraints, it is crucial to foster a culture of maintenance for malaria elimination and potential eradication. Our findings indicate that achieving malaria elimination is associated with a high level of utilization and consistent funding of interventions. The work presented in this paper can potentially contribute to developing effective strategies for malaria control and elimination. By identifying key transmission pathways and emphasizing the importance of intervention maintenance, our findings can guide decision-makers and stakeholders in their efforts to combat malaria and improve public health.

## 6 Author Contributions

M. A. Korsah, S. T. Johnston, J. A. Flegg and C. R. Walker conceived the study. M. A. Korsah conducted the simulations and drafted the manuscript. K. E. Tiedje and K. P. Day provided feedback on the modelling of the asymptomatic class and the malaria control interventions considered in the study. S. T. Johnston, K. E. Tiedje and K. P. Day critically reviewed and revised the manuscript for intellectual content. J. A. Flegg, and C. R. Walker oversaw project coordination and contributed to the manuscript revisions. All authors read and approved the final manuscript.

## 7. Acknowledgments

We gratefully acknowledge the support of the Melbourne Research Scholarship, the National Institute of Allergy and Infectious Diseases and the National Institutes of Health for their financial assistance, enabling the completion of this research.

## 9 Data Availability

Data sharing does not apply to this paper, as there were no generated or analyzed data sets during the study.

### A Model Analysis and Simulation Results

#### A.1 Basic reproduction number using the next generation method

We start by finding the new infection matrix:

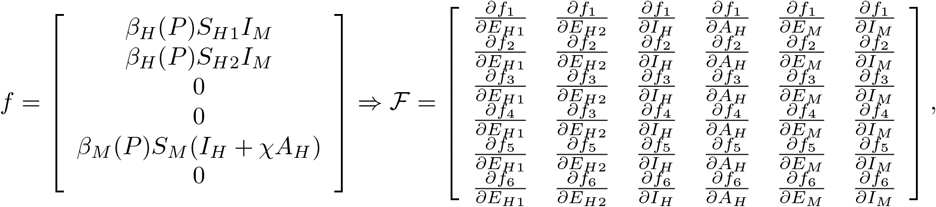

which gives

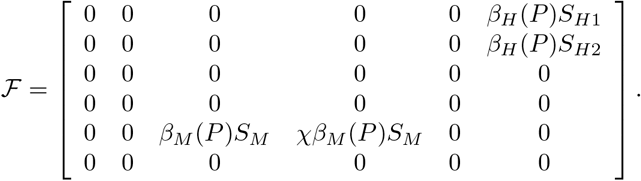

And the transition matrix:

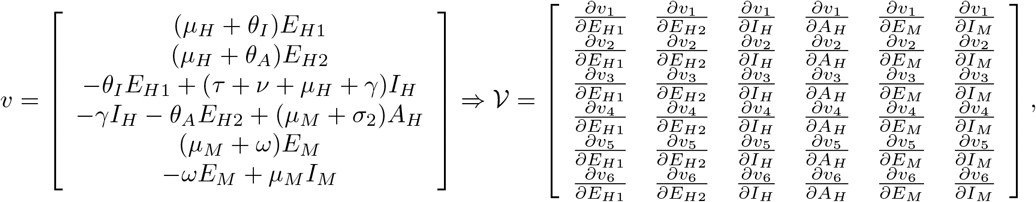

which gives

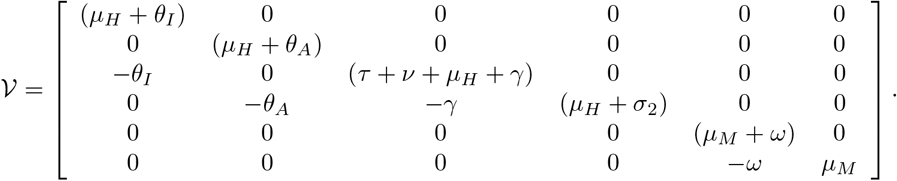

We compute ℛ_0_ by finding the spectral radius, *ρ* of the matrix product of *ℱ 𝒱* ^−1^ where

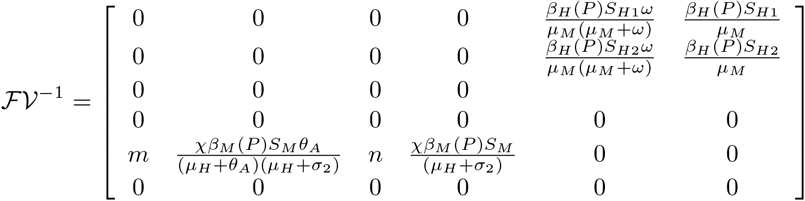

and

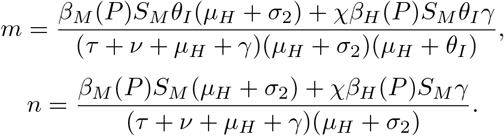

We consider the model at the disease-free equilibrium (DFE),

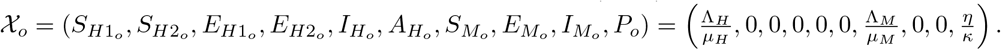

Note that for the DFE, 𝒳_*o*_, 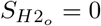 since *S*_*H*2_ represents individuals with partial immunity derived from an earlier infection. We therefore arrive at:

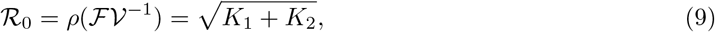

where *K*_1_ and *K*_2_ are defined as;

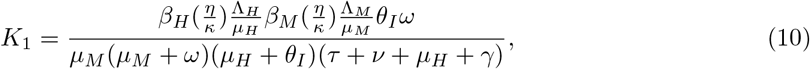

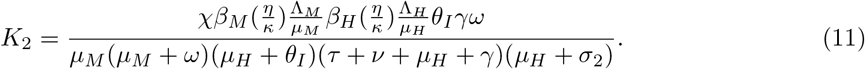

#### A.2 Example simulation results of the model

We provide numerical simulations of the model framework utilizing parameter estimates from previous literature [30, 39, 40, 41] presented in Table 1 to generate graphical representations of the model dynamics towards the disease-free (DFE) and endemic equilibria (EE). In exploring the model system at the two equilibria, the basic reproduction number, ℛ_0_ was set at ℛ_0_ *<* 1 and ℛ_0_ *>* 1 to depict the DFE and EE behavioural trends. We present the trends of the system in both the host and vector populations.

From our example simulation in Figure 6a, showing the disease-free equilibrium, it is noticed that the second class of susceptible individuals, *S*_*H*2_, approaches zero like the diseased classes of the human population as ℛ_0_ *<* 1. This behaviour is contrary to the *S*_*H*1_ class that approaches the total human population size 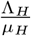. When one reconciles the behaviour with the description of the sub-classes, the disparity in the behaviours of the two susceptible sub-classes becomes clear; uninfected, non-immune humans make up the first susceptible class, whereas individuals in the second class are uninfected but partially immune, having acquired this immunity from a prior infection. Thus by inferring from the model diagram, Figure 1, as *I*_*H*_ decreases towards the DFE, *S*_*H*2_ will decrease as well.

**Figure 6.**
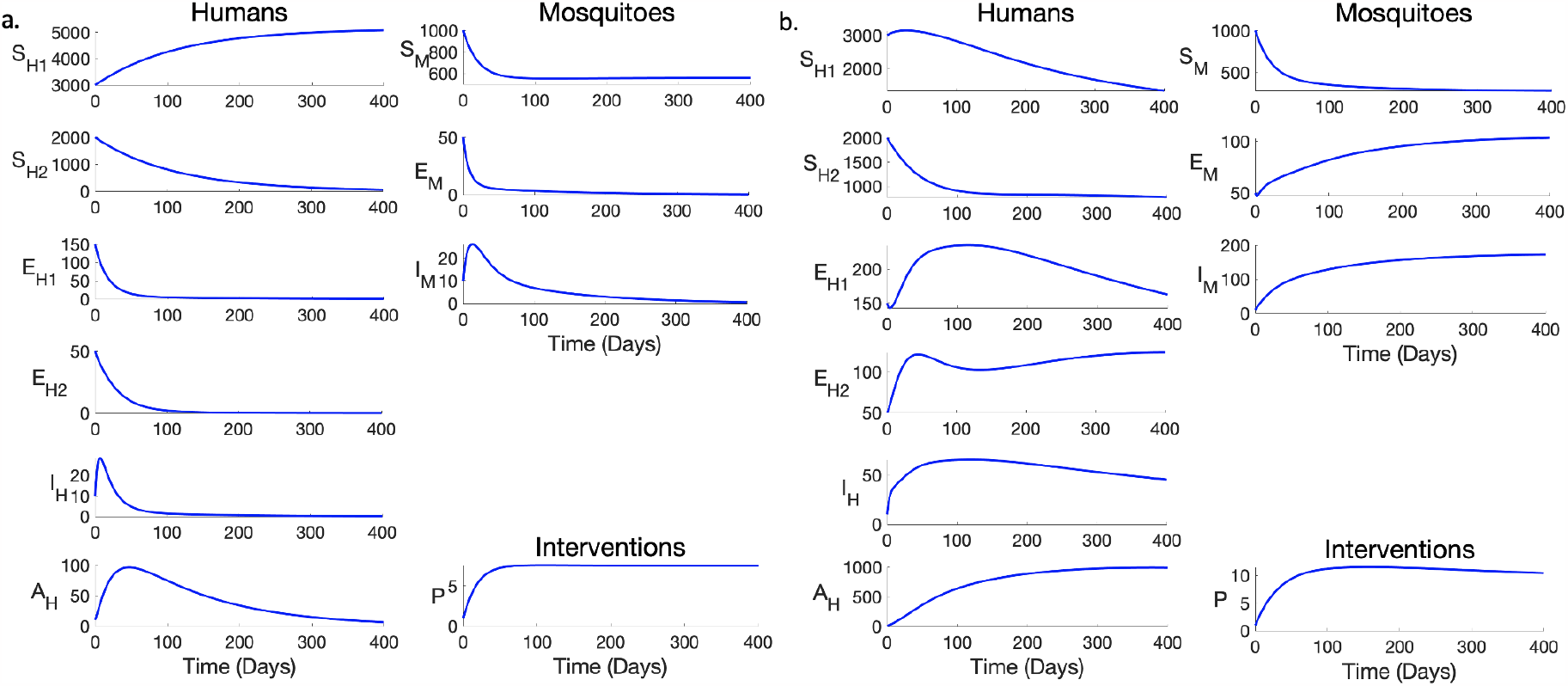
Example model simulation results, showing human and mosquito population dynamics. a.Example simulation result of the model at ℛ_0_ = 0.35. b. Simulation result of the model at ℛ_0_ = 2.26. The baseline values given in Table 1 are employed as parameter values for the ℛ_0_ *<* 1 simulation result, while setting *δ* = 3, *ψ*_*H*_ = 0.5 and *ψ*_*M*_ = 0.5 for the ℛ_0_ *>* 1 simulation. The initial conditions was set at (*S*_*H*1_(0), *S*_*H*2_(0), *E*_*H*1_(0), *E*_*H*2_(0), *I*_*H*_ (0), *A*_*H*_ (0), *S*_*M*_ (0), *E*_*M*_ (0), *I*_*M*_ (0), *P* (0)) = (3000, 2000, 150, 50, 10, 10, 1000, 50, 10, 1).

In Figure 6b, where ℛ_0_ *>* 1, we observe a rise in the diseased classes of the model which shows that an increase in the number of infected mosquitoes has a corresponding effect on the number of infected humans and vice versa. In the susceptible curves however, there is sharp decline resulting from the dynamics in the infected compartment, and later an increase as individuals recover and as the model solution progresses towards equilibrium. The ripple effect of infection is supported here by the graphical results of the diseased and susceptible classes in Figure 6b where an increase or decrease in the diseased classes of mosquitoes has a corresponding effect on the diseased classes of humans and vice versa.

